# Emergency Physician Ultrasound-guided Nerve Block Training Simulation Assessment: a Prospective Cohort Study

**DOI:** 10.64898/2026.02.06.26345652

**Authors:** Daniel Mercader, Reginald Lerebours, Erica Peethumnongsin, Catherine Staton, Maragatha Kuchibhatla, Rebecca Theophanous

**Affiliations:** Duke University School of Medicine, Department of Emergency Medicine, Durham, NC 27710; Duke University School of Medicine, Department of Biostatistics and Bioinformatics, Durham, NC 27710; Duke University Global Health Institute, Durham, NC 27710; Durham Veterans Affairs Healthcare System, Durham, NC, 27710

**Keywords:** serratus anterior plane block, ultrasound-guided nerve blocks, emergency department nerve blocks, point-of-care ultrasound, POCUS training

## Abstract

**Background:** Standardized training and competency testing is needed for appropriate point-of-care ultrasound (POCUS) clinical use. Our study objective assesses a low-fidelity simulation pig model workshop and tests the knowledge and technical skills of emergency medicine (EM) clinicians when performing simulated ultrasound-guided serratus anterior nerve block (UG-SANB).

**Methods:** EM residents, attendings, and advanced practice providers (APP) participated in a prospective cohort study, completing a one-time simulation-based UG-NB training session at a single academic medical center between November 2024 to February 2025. Training model acceptability, appropriateness, and feasibility was assessed using the validated AIM-IAM-FIM tool (pre/post-surveys). Effectiveness outcomes were participant knowledge score, technical skill score, and self-rated confidence in performing NBs pre-, post-, and 3-months post-intervention. Clinical ED-performed ultrasound-guided nerve blocks were reported pre-/post-intervention. Scores were summarized using mean (S.D.) and total question percent correct. Paired individual assessments were compared pre/post-intervention using paired t-tests and group assessments using t-tests for normal data distribution.

**Results:** 63/104 ED providers (60.6%) responded to surveys pre-intervention and 57 post-intervention (54.8%). 63 providers (16 EM attendings, 33 residents, and 14 APPs) underwent SANB training and testing. Participant survey responses reported the training model was acceptable, appropriate, and feasible (at least 54/57 agreed or strongly agreed for all three). Mean knowledge scores were 85% (SD 14.8%) post- and 70% (SD 18.2%) 3-months post-workshop. Mean technical skills exam scores were 98% (SD 4.5%) post- and 95% (5.8%) 3-months post-intervention. Perceived confidence in teaching clinical NBs increased pre-/post-intervention (from 11.3% to 58.2%) and for SANB (3.2% to 70.2%). Clinically performed NBs at pre and post were 21 and 15 respectively.

**Conclusion:** Emergency clinicians’ knowledge, technical skills, and confidence scores increased after an UG-NB training intervention. This standardized, reproducible simulation model could improve clinical skills and patient care outcomes but needs additional steps to increase clinical UG-NB performance.

## Introduction

### Background

Maintaining procedural skills competency is an important aspect of medical education and physician training, especially in emergency medicine (EM).^1–4^ A standardized assessment and training model for EM procedural skills does not exist, and achieving knowledge and skill sustainability is difficult. Simulation tools are a critical hands-on training component with real-time equipment in a controlled environment, including for learning point-of-care ultrasound (POCUS).^5–6^ POCUS is an important bedside tool used by emergency physicians in medical decision making and critical procedural skills such as intravenous access or nerve blocks for pain management.^7–8^ Currently, there is no reproducible, standardized procedural skill training model for ultrasound-guided nerve blocks (UG-NB) for EM physicians.

### Importance

Prior studies have investigated simulation training for UG-NB performance such as femoral nerve, intercostal nerve, and serratus anterior plane nerve block (SANB) with EM residents.^6,9–11^ However, the literature is scarce for evaluation and training of attending physicians who have been working clinically for years and already have an established practice pattern.^12–13^ Emphasizing continued faculty education is important for maintaining practical skills, particularly for those at academic centers.^14^

UG-SANB can be performed safely with proper training.^15–16^ As rib fractures are the most common injury in blunt trauma (10-38% of ED blunt trauma patients), UG-SANB can have a large impact in improving patient outcomes.^16–17^ Thoracic NB performed by anesthesiologists in the inpatient setting are safe and effective in achieving pain control and reducing significant complications including pneumonia, prolonged hospitalizations, and death.^15–16^ More recent studies have shown that NB performance by EM physicians similarly provides good pain control with very low complication rates.^17–19^ Nevertheless, the invasive nature of the procedure and possibility for pneumothorax, internal bleeding, or other complications necessitates training. Thus, EM physicians still feel uncomfortable performing or supervising residents for these procedures at hospitals nationwide, including major trauma centers.^17–19^

### Goals of this investigation

Our primary aim was to assess EM physician and APP knowledge and technical skill in performance of ultrasound-guided SANB in a low-fidelity simulation model workshop. We also evaluated the acceptability, appropriateness, and feasibility of our EM physician education and simulation workshop implementation strategy using the AIM-IAM-FIM tool.^20^ By developing and evaluating an EM physician training program to safely perform and teach ultrasound-guided procedures, we anticipate our didactic training program will be able to improve physician skills and thereby patient outcomes nationwide.

## Methods

### Study design and setting

This was a prospective cohort study including 56 attending physicians, 36 EM resident physicians (PGY1-3), and 26 APPs at a single academic medical center with 1,048 inpatient beds, level 1 trauma center designation, and over 85,000 ED visits per year. Exclusion criteria are ED nursing and ED technicians, as their ultrasound training needs differ from physicians and APPs. All APPs employed at our site’s ED were physician assistants during the study period. Simulation workshops were held from November 2024 to February 2025.

### Selection of participants

Our sampling method was convenience sampling with recruitment through emails and monthly EM faculty meetings or weekly EM resident conference.

Participants gave implied consent by reading the emailed study data sheet, completing the pre-assessment survey, and participating in the workshop. They could withdraw from the study at any time. We followed the Strengthening the Reporting of Observational Studies in Epidemiology (STROBE) guidelines for reporting on prospective cohort studies.^21^(**Supplemental File 1**) The study was performed with Duke University Health System Institutional Review Board (IRB) approval and implied informed consent was obtained via participant review of study materials per the IRB protocol (Pro00114989). The study met the institutional ethical standards and followed the Declaration of Helsinki.

### Interventions/Study procedures

a) EM providers participated in a survey and written knowledge assessment pre/post-intervention and 3-months post-intervention consisting of multiple-choice questions about serratus anterior nerve blocks (UG-SANB) to assess feasibility. Participants reviewed online pre-reading material and a video (*Free Open Access Meducation from the Core Ultrasound and Academy of Emergency Ultrasound websites*) for self-learning that was emailed to providers at least one week before the training workshop. Subjects participated in an in-person training session at the hospital’s simulation center between November 2024 to February 2025.

The one-hour workshop involved a 15-minute introductory lecture given by ultrasound faculty with an overview on NBs, SANB anatomy and example images, and identifying structures. Then participants actively practiced UG-SANB on a low-fidelity pork belly simulation model with ultrasound faculty guidance.(**Supplemental File 2**)

### Measurements

Knowledge and technical skills scores and survey data were collected via the institution’s secured REDCap server. Clinical data was collected from the Butterfly Enterprise POCUS archiving software for ED-performed nerve blocks (quality assurance image review scores and number of clinical POCUS NBs performed pre-/post-intervention).

a. EM providers individually completed an observed structured clinical examination (OSCE) technical skills assessment using the same simulation model with a single ultrasound-trained rater using a validated learning checklist (Modified Cheung checklist, 15-items).(**Supplemental File 3**) The assessment was proctored by the same ultrasound faculty rater each time in a secured area without other people present to avoid bias and maintain confidentiality. Image acquisition was tested with structure recognition on a normal healthy human model. Participants also completed an 11-question knowledge assessment and a post-survey created by the study primary investigator based on surveys validated in other studies and piloted with a physician from another institution not participating in the study.
b. To evaluate our simulation workshop implementation strategy, we used the Acceptability of Intervention Measure (AIM), Intervention Appropriateness Measure (IAM), and Feasibility of Intervention Measure (FIM).^20^ AIM-IAM-FIM is a reliable, validated tool that can be used to monitor and evaluate the success of implementation efforts.^20^ All values were documented via participant survey responses using a Likert scale. EM physicians completed a post-intervention survey to evaluate the implementation of our simulation workshop. The survey questions are based on the AIM-IAM-FIM implementation outcome measures to assess acceptability, appropriateness, and feasibility of the simulation workshop. ^20^ Post-workshop survey questions assessed the simulation workshop implementation: 1) **acceptability** –participant appeal, likability, and approval of the simulation workshop and teaching format, 2) **appropriateness** – whether participants feel the simulation workshop is fitting, suitable, applicable, and appropriate for their training level and applicable to clinical use, and 3) **feasibility** – whether the simulation workshop is implementable, possible to complete in the given timeframe, doable to attend and participate, and ease of using the simulation model and completing the assessments.(**Supplemental File 4**)^20^

### Outcomes

Individual providers were scored via RedCap surveys for knowledge and technical skills assessments. Demographic participant data was compared between EM attending, EM resident, and APP groups and between training levels (PGY1-3, fellow, attending). Pre/post-intervention survey responses regarding comfort and confidence in performing and teaching UG-SANB, participants’ clinical experience performing NB, and a workshop evaluation assessed acceptability, appropriateness, and feasibility. We collected de-identified clinical data on the number of SANB performed.

### Statistical analysis plan

a) Outcomes included pre-/post-intervention knowledge score, technical skill score, and self-rated comfort in performing nerve blocks. Competency or pass rate was defined as >90% correct on the knowledge assessment and technical skills exam. Scores were summarized using mean with standard deviations (S.D.), median with interquartile range (IQR), and total question percent correct.

Paired within physician individual assessments of changes in assessment scores of a) pre/post and b) pre- to 3-months post-workshop were compared using paired t-tests or Wilcoxon tests depending on whether the change scores were normally or non-normally distributed. We report the percentage correct of steps performed on the validated skills checklist, as measured by direct observation by an ultrasound-trained physician, for individual EM attending, resident, and APP technical skills for needle placement in a simulation model and image acquisition of correct identification of structures and presumed needle position on a normal human model.

These percentages were reported for each of the two samples along with 95% confidence interval, and the difference in proportions between the two samples were examined using a z-test. All these tests were reported with and without adjusting for multiple testing (i.e. multiple comparison of questions at pre/post and from pre- to 3 months post-workshop, in addition to the summary score of these questions).

Written knowledge assessment percent correct between EM attending vs EM resident and EM physician vs APP groups and training levels (PGY1-3, attending, APP) was compared using Chi-square tests. Scores >7 (75% percent correct) were considered as passing the assessment. The satisfaction scales for group assessments were performed using two-sample t-tests or Wilcoxon Rank tests depending on data distribution. Physician confidence and comfort with performing and teaching UG-SANB per survey responses (Likert scales) were summarized via mean (S.D.), median (IQR), change scores, and 95% confidence intervals.

b) Simulation workshop implementation acceptability, appropriateness, and feasibility per post-intervention survey responses (Likert scales) was summarized via mean (S.D.), median (IQR), and 95% confidence intervals.

### Sample size and power

For EM attendings, using a two-sided, paired-difference t-test, Type I error rate (α) of 0.05, and assumed standard deviation of 2, with a sample size of 56 pairs from a population of 10,000 pairs, to detect a paired mean difference of 2, the power is 1. For residents, with a sample size of 36 pairs and the same values, the power is 0.99995. A parallel two-group design will be used to test the difference between two means. Using a two-sided, two-sample equal-variance t-test, based on our expected population effect size of 0.8, Type I error rate (α) of 0.05, and sample size of 56 in Group 1 and 36 in Group 2, the power is 0.99. With an effect size of 0.63, the power is 0.65.(PASS 2023, version 23.0.1)^22^

## Results

### Characteristics of study subjects

63/104 ED providers (60.6%) responded to surveys pre-intervention and 57 post-intervention (54.8%). There were 63 providers (16 EM attendings, 33 residents, and 14 APPs) who underwent SANB training and testing from November 2024 to February 2025. Participant demographics are listed in Supplemental File 5.

### Main results

#### Knowledge Skill Tests (Table 1, Figure 1)

There are 63 unique participants (APP=14, Attending=16, Resident=33) corresponding to 94 observations for the “study knowledge” assessment (31 participants have two timepoints). The mean percentage score was 84% (SD=14.8%) and the median 91% (IQR: 73%, 91%) post-intervention. At 3-months post-intervention, the mean percentage correct score was 70% (18.2%) and median 73% (IQR: 64%, 82%).

**Figure 1:**
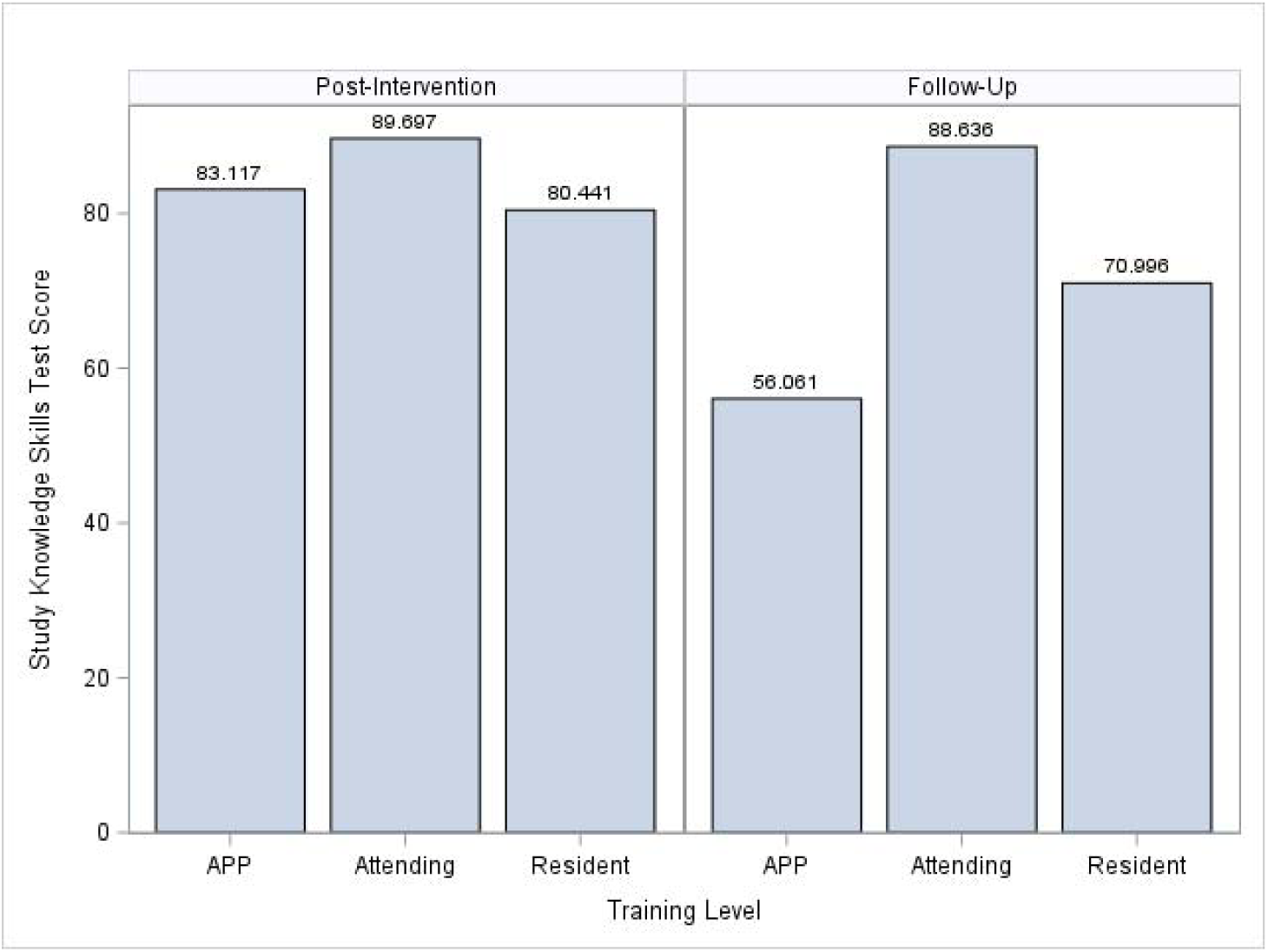
Study knowledge test percent score by group at post-intervention and 3-months post-intervention (APP=advanced practice provider)

**Table 1:** Study Knowledge results (percent score) by group at post- and 3-months-post intervention.

|  | APP<br>(N=14) | Attending<br>(N=17) | Resident<br>(N=33) | Total<br>(N=64) | p value |
| --- | --- | --- | --- | --- | --- |
| **Study Knowledge Skill Test Percent Score (post-intervention)** |  |  |  |  | 0.02<sup>1</sup> |
| N | 14 | 16 | 33 | 63 |  |
| Mean (SD) | 83 (12.8) | 90 (13.9) | 80 (15.3) | 84 (14.8) |  |
| Median (IQR) | 91 (73, 91) | 91 (91, 100) | 82 (73, 91) | 91 (73, 91) |  |
| Range | (55-100) | (45-100) | (45-100) | (45-100) |  |
| **Study Knowledge Skill Test Score passed (>75%) (post-intervention)** | 10 (71.4%) | 14 (87.5%) | 22 (66.7%) | 46 (73.0%) | 0.31<sup>2</sup> |
| N | 14 | 16 | 33 | 63 |  |
| **Study Knowledge Skill Test Score (3-months post-intervention) (N=31)** |  |  |  |  | 0.009<sup>1</sup> |
| Mean (SD) | 56 (13.4) | 89 (8.7) | 71 (17.6) | 70 (18.2) |  |
| Median (IQR) | 59 (45, 64) | 86 (82, 95) | 73 (64, 82) | 73 (64, 82) |  |
| Range | (36-73) | (82-100) | (36-100) | (36-100) |  |
| **Study Knowledge Skill Test Score passed (>75%) (3-months post-intervention) (N=31)** | 0 (0.0%) | 4 (100.0%) | 9 (42.9%) | 13 (41.9%) | 0.005<sup>2</sup> |

|  | APP<br>(N=14) | Attending<br>(N=17) | Resident<br>(N=33) | Total<br>(N=64) | p value |
| --- | --- | --- | --- | --- | --- |
| **Study Knowledge Skill Test (post-intervention vs 3-month follow-up)** |  |  |  |  | 0.14<sup>1</sup> |
| N | 6 | 4 | 21 | 31 |  |
| Mean (SD) | -24 (18.8) | -7 (8.7) | -10 (17.9) | -12 (17.8) |  |
| Median (IQR) | -18 (-36, -9) | -5 (-14, 0) | -9 (-18, 0) | -9 (-18, 0) |  |
| Range | (-55--9) | (-18-0) | (-55-18) | (-55-18) |  |

<sup>1</sup>Kruskal Wallis <sup>2</sup>Fisher Exact**Table 2: Technical Skills test percent scores by group at post- and 3-months post-intervention**

Of the 31 participants that have two assessment points (APP=6, Attending=4, Resident=21), the mean percentage score was 83% (SD=15.3%) and median 91% (IQR: 73, 91%) post-intervention. At 3-months post-intervention, the mean percentage score was 70% (SD=18.2) and median 73% (IQR: 64, 82%).

On average, participants scored 12% lower on their follow-up assessment compared to their post-intervention assessment. 73% of participants (46/63, 1 missing) had a “study knowledge” score of greater than 75% at post-intervention, and 42% of participants (13/31) had a score of greater than 75% at the 3-months post-intervention follow-up.

#### Technical Skill Tests (Table 2, Figure 2)

There are 86 unique participants (APP=17, Attending=19, Resident=48, 2 not reported) corresponding to 104 observations for the “technical skills assessment (18 participants had two time points). The mean percentage correct score for the “technical skills” assessment across all participants (n=16) is 98% (SD=4.5%) and median 100% (IQR: 93%, 100%) post-intervention. At 3-months post-intervention, the mean percentage correct score is 95% (SD=5.8%) and median 97% (IQR: 93%, 100%).

**Figure 2:**
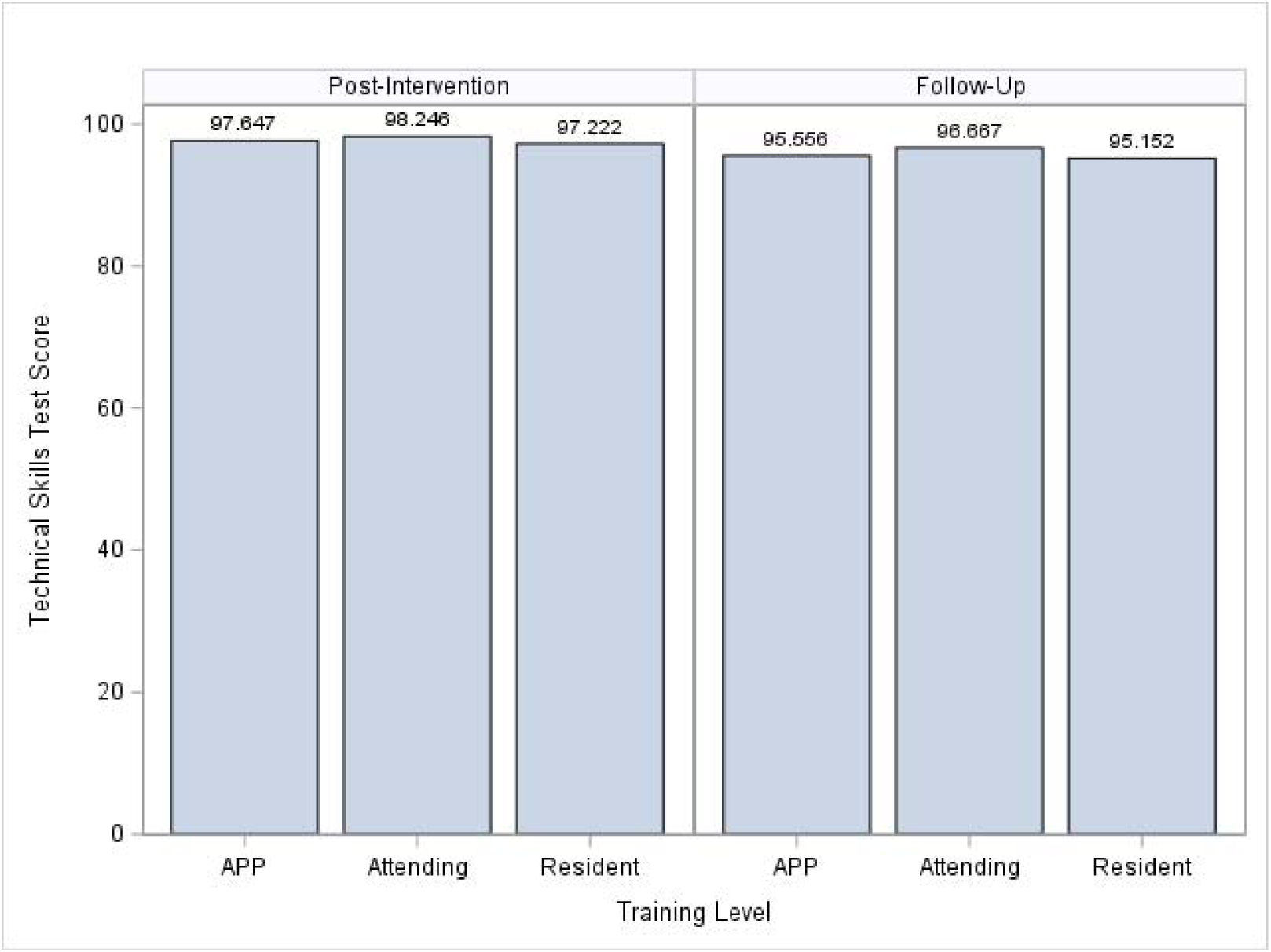
Technical skills test percent score by group at post-intervention and 3-months post-intervention (APP=advanced practice provider)

**Table 2:**
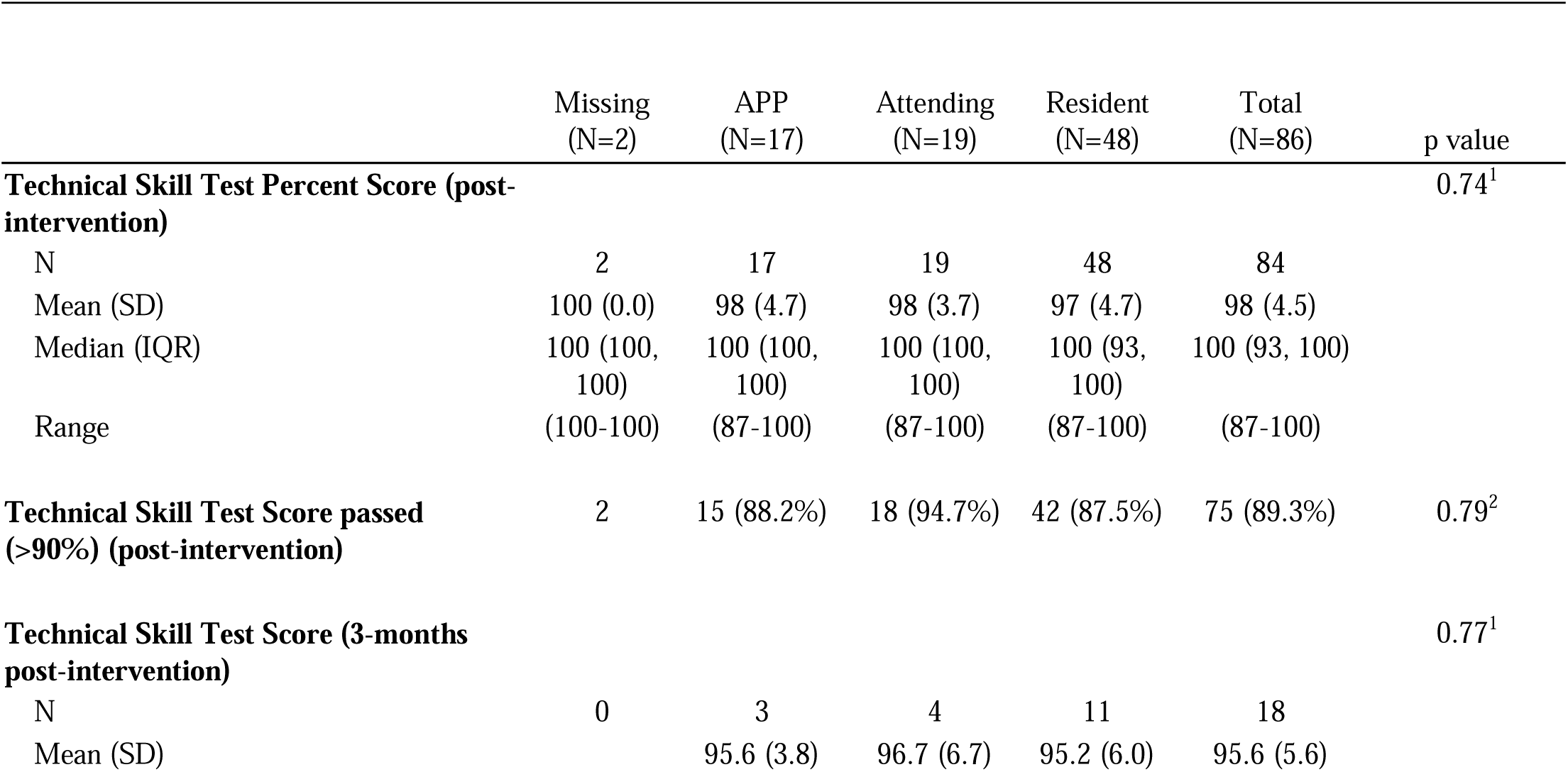
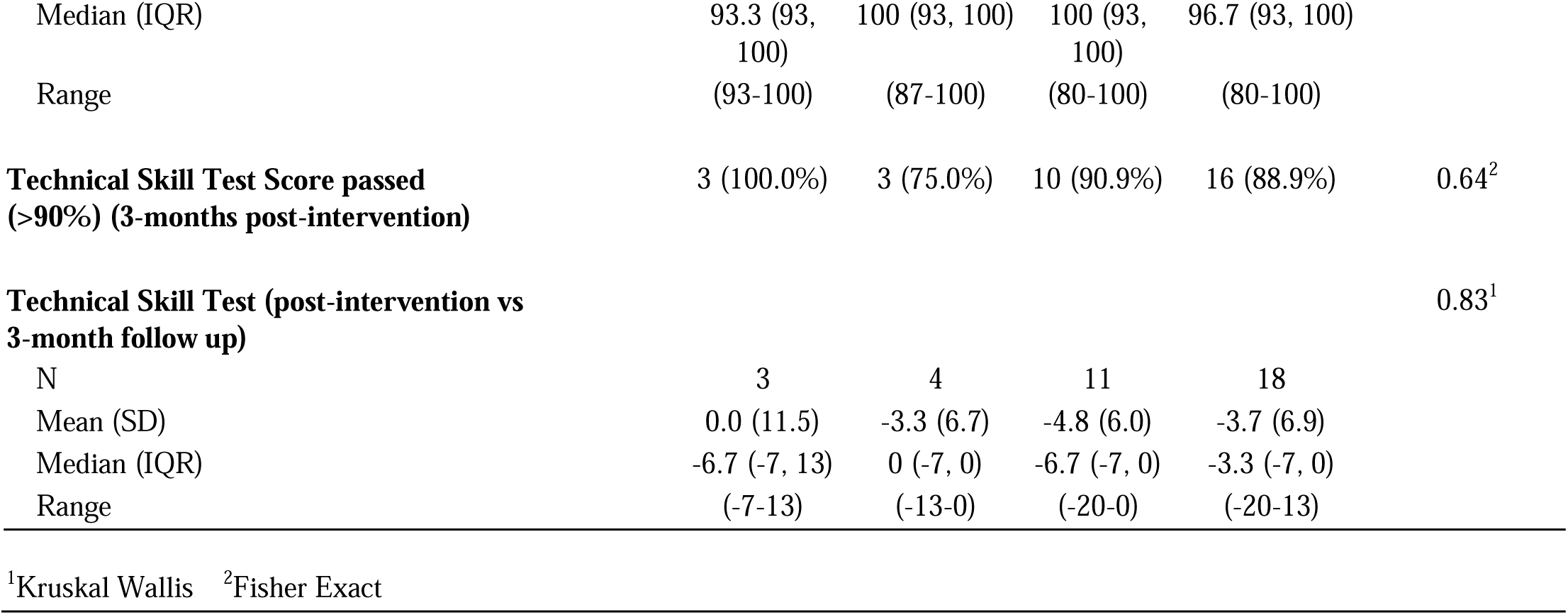
Technical Skills test percent scores by group at post- and 3-months post-intervention.

Of the 18 participants that have two assessment points (APP=2, Attending=3, Resident=11, 2 not reported), the mean percentage correct score was 99% (SD=3.1%) and median 100% (IQR: 100%, 100%) post-intervention. At 3-months post-intervention, the mean percentage correct score was 96% (SD=5.6) and median 97% (IQR: 93%, 100%).

On average, participants scored 3.7% lower on their 3-month follow-up assessment compared to their post-intervention assessment. 89.3% of participants (75/84, 2 missing) had a “technical skills” score of greater than 90% post-intervention and 88.9% of participants (16/18) had a score of greater than 90% at the 3-month follow-up.

#### Self-confidence tests (from Pre-Post survey assessments)

Before the intervention, 11.3% of participants (7/62, 1 missing) felt either “confident” or “very confident” in teaching others how to perform any clinical nerve block with ultrasound-guidance. 3.2% of participants (2/62, 1 missing) felt either “confident or “very confident” in performing serratus anterior clinical nerve block with ultrasound-guidance. Similarly, 3.2% of participants (2/62, 1 missing) felt either “confident or “very confident” in teaching others to perform UG-SANB. All 63 participants either “agree” or “strongly agree” that POCUS is a useful tool for medical decision making in the ED.(**Table 3a**)

**Table 3a:** Pre-survey point-of-care ultrasound emergency provider confidence assessment.

|  | Total<br>(N=63) |
| --- | --- |
| **How confident do you feel teaching others how to perform ANY clinical nerve block with ultrasound-guidance before the intervention?** |  |
| Missing | 1 |
| Very unconfident | 29 (46.8%) |
| unconfident | 11 (17.7%) |
| neutral | 15 (24.2%) |
| confident | 6 (9.7%) |
| very confident | 1 (1.6%) |
| **How confident do you feel performing serratus anterior clinical nerve block with ultrasound-guidance before the intervention?** |  |
| Missing | 1 |
| Very unconfident | 30 (48.4%) |
| unconfident | 21 (33.9%) |
| neutral | 9 (14.5%) |
| confident | 1 (1.6%) |
| very confident | 1 (1.6%) |
| **How confident do you feel teaching others how to perform serratus anterior clinical nerve block with ultrasound-guidance before the intervention?** |  |
| Missing | 1 |
| Very unconfident | 38 (61.3%) |
| unconfident | 14 (22.6%) |
| neutral | 8 (12.9%) |
| confident | 1 (1.6%) |
| very confident | 1 (1.6%) |
| **Do you believe that POCUS is a useful tool for medical decision making in the ED?** |  |
| agree | 13 (20.6%) |
| strongly agree | 50 (79.4%) |

After the intervention, 58.2% of participants (32/55, 3 missing) felt either “confident” or “very confident” in teaching others how to perform any clinical nerve block with ultrasound-guidance. 84.2% of participants (48/57, 1 missing) felt either “confident or “very confident” in performing UG-SANB. 70.2% of participants (40/57, 1 missing) felt either “confident or “very confident” in teaching others to perform serratus anterior clinical nerve block with ultrasound-guidance. 98.2% of participants (56/57, 1 missing) either “agree” or “strongly agree” that POCUS is a useful tool for medical decision making in the ED. (**Table 3b**)

**Table 3b:** Post-survey point-of-care ultrasound emergency provider confidence Assessment.

|  | Total<br>(N=58) |
| --- | --- |
| **How confident do you feel teaching others how to perform ANY clinical nerve block with ultrasound-guidance after the intervention?** |  |
| Missing | 3 |
| Very unconfident | 3 (5.5%) |
| unconfident | 3 (5.5%) |
| neutral | 17 (30.9%) |
| confident | 27 (49.1%) |
| very confident | 5 (9.1%) |
| **How confident do you feel performing serratus anterior clinical nerve block with ultrasound-guidance after the intervention?** |  |
| Missing | 1 |
| Very unconfident | 1 (1.8%) |
| unconfident | 2 (3.5%) |
| neutral | 6 (10.5%) |
| confident | 38 (66.7%) |
| very confident | 10 (17.5%) |
| **How confident do you feel teaching others how to perform serratus anterior clinical nerve block with ultrasound-guidance before the intervention?** |  |
| Missing | 1 |
| Very unconfident | 3 (5.3%) |
| unconfident | 2 (3.5%) |
| neutral | 12 (21.1%) |
| confident | 31 (54.4%) |
| very confident | 9 (15.8%) |
| **Do you believe that POCUS is a useful tool for medical decision making in the ED?** |  |
| Missing | 1 |
| Strongly disagree | 1 (1.8%) |
| agree | 10 (17.5%) |
| strongly agree | 46 (80.7%) |

The AIM-IAM-FIM post-survey results are reported in **Table 4a/4b**. Participant survey responses stated the training model was acceptable, appropriate, and feasible (at least 54/57 respondents agreed or strongly agreed for all three). Overall, people were satisfied with the course (56/57 agreed or strongly agreed) and 54/57 reported that they would apply the knowledge and skills from the workshop to teach others. Qualitative feedback from participant comments stated that “the hands-on aspect of the workshop” was most useful to them (22/57 responses), as well as “being able to practice tracking the needle with the ultrasound” and “the simulation model” (9 responses each).

**Table 4a:** Post-Intervention AIM-IAM-FIM Survey Course Assessment (Part 1)

|  | Total<br>(N=58) |
| --- | --- |
| **I approve of the education and simulation workshop**<br>**(acceptability)** |  |
| Missing | 2 |
| Strongly disagree | 1 (1.8%) |
| agree | 9 (16.1%) |
| strongly agree | 46 (82.1%) |
| **The simulation workshop teaching format is appealing**<br>**to me (acceptability)** |  |
| Missing | 1 |
| Strongly disagree | 1 (1.8%) |
| agree | 9 (15.8%) |
| strongly agree | 47 (82.5%) |
| **I like the education and simulation workshop**<br>**timeframe (acceptability)** |  |
| Missing | 2 |
| Strongly disagree | 1 (1.8%) |
| disagree | 1 (1.8%) |
| agree | 11 (19.6%) |
| strongly agree | 43 (76.8%) |
| **I like the hands-on aspect of the simulation model**<br>**(acceptability)** |  |
| Missing | 1 |
| Strongly disagree | 1 (1.8%) |
| agree | 9 (15.8%) |
| strongly agree | 47 (82.5%) |
| **The simulation workshop structure and format are**<br>**fitting (appropriateness)** |  |
| Missing | 2 |
| Strongly disagree | 1 (1.8%) |
| agree | 10 (17.9%) |
| strongly agree | 45 (80.4%) |
| **The simulation workshop seems suitable to my**<br>**training level (appropriateness)** |  |
| Missing | 1 |
| Strongly disagree | 1 (1.8%) |
| agree | 10 (17.5%) |
| strongly agree | 46 (80.7%) |

**Table 4a: Post-Intervention AIM-IAM-FIM Survey Course Assessment (Part 1)**
|  | Total<br>(N=58) |
| --- | --- |
| **The simulation workshop seems applicable for use in clinical care (appropriateness)** |  |
| Missing | 1 |
| Strongly disagree | 1 (1.8%) |
| agree | 11 (19.3%) |
| strongly agree | 45 (78.9%) |
| **The education and simulation workshop are a good match as a simple and adaptable model for ultrasound procedural training (appropriateness)** |  |
| Missing | 1 |
| Strongly disagree | 1 (1.8%) |
| agree | 10 (17.5%) |
| strongly agree | 46 (80.7%) |
| **The simulation workshop and assessments seem implementable and reproducible for other procedures (feasibility)** |  |
| Missing | 1 |
| Strongly disagree | 1 (1.8%) |
| agree | 11 (19.3%) |
| strongly agree | 45 (78.9%) |
| **The simulation workshop is possible to complete in the given time (feasibility)** |  |
| Missing | 1 |
| Strongly disagree | 1 (1.8%) |
| agree | 11 (19.3%) |
| strongly agree | 45 (78.9%) |
| **It is doable to attend and participate in the simulation workshop (feasibility)** |  |
| Missing | 1 |
| Strongly disagree | 1 (1.8%) |
| neutral | 1 (1.8%) |
| agree | 12 (21.1%) |
| strongly agree | 43 (75.4%) |
| **The simulation model seems easy to use. (feasibility)** |  |
| Missing | 1 |
| Strongly disagree | 1 (1.8%) |

**Table 4a: Post-Intervention AIM-IAM-FIM Survey Course Assessment (Part 1)**
|  | Total<br>(N=58) |
| --- | --- |
| neutral | 1 (1.8%) |
| agree | 11 (19.3%) |
| strongly agree | 44 (77.2%) |

**Table 4b:** Post-Intervention Survey Course Assessment (Part 2)

|  | Total<br>(N=58) |
| --- | --- |
| **The pre-workshop self-learning training materials assisted my learning.** |  |
| Missing | 2 |
| Strongly disagree | 1 (1.8%) |
| neutral | 7 (12.5%) |
| agree | 21 (37.5%) |
| strongly agree | 27 (48.2%) |
| **The workshop teaching format was effective.** |  |
| Missing |  |
| Strongly disagree | 1 (1.8%) |
| agree | 11 (19.3%) |
| strongly agree | 45 (78.9%) |
| **The content covered was appropriate to my skill level.** |  |
| Missing | 1 |
| Strongly disagree | 1 (1.8%) |
| agree | 11 (19.3%) |
| strongly agree | 45 (78.9%) |
| **I gained new knowledge and skills by participating in the workshop.** |  |
| Missing | 1 |
| Strongly disagree | 1 (1.8%) |
| agree | 12 (21.1%) |
| strongly agree | 44 (77.2%) |

**Table 4b: Post-Intervention Survey Course Assessment (Part 2)**
|  | Total<br>(N=58) |
| --- | --- |
| **I will apply the knowledge and skills from the workshop for clinical patient care.** |  |
| Missing | 1 |
| Strongly disagree | 1 (1.8%) |
| agree | 12 (21.1%) |
| strongly agree | 44 (77.2%) |
| **I will apply the knowledge and skills from the workshop to teach other learners.** |  |
| Missing | 2 |
| Strongly disagree | 1 (1.8%) |
| neutral | 1 (1.8%) |
| agree | 12 (21.4%) |
| strongly agree | 42 (75.0%) |
| **Overall, I am satisfied with the course.** |  |
| Missing | 1 |
| Strongly disagree | 1 (1.8%) |
| agree | 9 (15.8%) |
| strongly agree | 47 (82.5%) |
| **I would recommend this course to a colleague.** |  |
| Missing | 2 |
| Strongly disagree | 1 (1.8%) |
| agree | 10 (17.9%) |
| strongly agree | 45 (80.4%) |

Twenty-one clinical NBs were performed in the ED in the six months pre-intervention and 15 post-intervention. Since this was a procedural POCUS study, the quality assurance image review scores were reported as a yes/no response, and all studies achieved “meets criteria for credentialing”.

## Discussion

Our study assessed a low-fidelity pork belly simulation training model for UG-SANB. We achieved an 84% mean knowledge skill score post-intervention (SD 14.8%) and 70% (SD 18.2%) at 3-months post-intervention in EM attending and resident physicians and APPs. Technical skills scores were 98% mean (SD 4.5%) post-intervention and 95% (SD 5.8%) at 3-months post-intervention. The maintained higher scores suggest knowledge and technical skill retention. Participants felt more “confident” or “very confident” in teaching others how to perform any clinical nerve block after the workshop (from 11.3% pre- to 58.2% post-intervention) and for SANB specifically (increased from 3.2% pre- to 70.2% post-intervention). These findings with additional survey results suggest that the SANB training model may be acceptable, appropriate, and feasible for participating ED physicians and APPs.

*Point-of-care ultrasound (POCUS) is an important topic requiring procedural competency in EM physicians, including procedural applications such as intravenous access or nerve blocks for pain management*. Anesthesia literature has shown great benefit in performing regional anesthesia for patients with thoracic trauma, including in those with rib fractures, which are the most prevalent injury in blunt thoracic trauma (10-38% of ED blunt trauma patients).^15–16^ Regional anesthesia performed by anesthesiologists in the inpatient setting have demonstrated good pain control with fewer complications resulting from poor inspiratory effort such as pneumonia, prolonged hospitalization, and death.^15–17^ Further studies on ED-physician performed thoracic nerve blocks including SANB have shown similar good pain management with good safety profile in appropriately trained physicians.^17–19^ However, EM and other medical specialty attending physicians who do not perform these procedures on a regular basis, and especially those working at hospitals where resident trainees often perform the procedures, may feel uncomfortable performing nerve blocks and are hesitant to show a lack of knowledge compared to peers.^12–13,23^ This was true in our baseline pre-survey assessments, with most providers feeling uncomfortable performing or teaching clinical NB in the department.

Various training models have been described in the literature but lack standardization and reproducibility across physician training levels, and they are often time intensive. We created a simple model with easily accessible online resources plus in-person training. In contrast, Anstey et al used staged learning for teaching Internal Medicine (IM) faculty POCUS, with intensive in-person educational supervision, limited observed practice, and independent practice pathways for POCUS competency training and clinical use.^24^ Another study by Restrepo in IM and EM physicians found that teaching POCUS in mid-to-late career faculty was challenging due to feelings of vulnerability in acquiring new skills, emphasizing comfort over competence, spatial learning techniques, and a “bolus/drip” method with an intensive introductory course followed by longitudinal small group POCUS scanning sessions.^12^ We found similar problems in recruiting attending physicians to participate in the workshop, despite their ultrasound credentialing status (16/56 attendings participated). Because training and credentialing for POCUS does not have a standardized process, this leads to variable physician knowledge and skills, and lack of training remains a significant barrier to POCUS use at hospitals nationwide.^7–8,23,25^

*Maintaining procedural competency is important yet challenging in a diverse group of emergency medicine physicians and APPs with varying training backgrounds and clinical settings*. Recent medical education literature defines physician competency as the ability to safely and effectively practice the necessary skills to care for patients as an EM physician.^1–2^ Competency is an evolving and dynamic component of faculty development, thus its integration into clinical care and training is critical. Studies often describe training of students and resident trainees who are surrounded by teachers in a daily learning cycle and supported throughout their training program.^1–3,26^ We found a similar trend with high participation from the EM resident trainees (33/36 participated), who were eager to learn a new ultrasound and procedural skill.

Attending physicians, in contrast to resident trainees, have varying training backgrounds, different years of clinical experience and practice setting, and differing roles as they advance their clinical or educational career. Groleau and Knowles note that adults are often self-directed learners with a rich reservoir of experience and internal motivators, making it difficult to motivate mid-to-late career physicians to seek out or participate in continuing education or learning activities compared to resident physicians.^27–28^ Although resident training and competency testing is a popular topic in medical education research, few studies address EM attending physician competency.

*To address the lack of a reliable method for assessing ED physician and APP procedural competency, our study focused on a single, high-yield POCUS application as a model to create a standardized and reproducible training method for generalizability to other ultrasound-guided procedures*. By emphasizing active hands-on learning in a safe, controlled environment using a simulation model, we utilized the strengths of a task-oriented, practice-based pathway identified by Smalley et al. for successful adult learning.^13^ We used validated medical education research evaluation tools based on the ultrasound-guided regional anesthesia (UGRA) educational guidelines published by the American Society of Regional Anesthesia and Pain Medicine.^29^ Knowledge was assessed with a post/3-months post-intervention written selected-response exam (multiple choice questions). We also performed directly observed technical skills feedback with the Modified Cheung checklist and image acquisition method that was validated in another nerve block study, achieving a sustained increase in both knowledge and technical skills scores (70% and 95% at 3-months post-intervention, respectively).^9,30–31^

We previously successfully implemented a comprehensive POCUS program at a single Veterans Affairs ED as a model for national dissemination. When comparing our results to that study, we found that EM attending physicians were enthusiastic about learning POCUS, with a four-fold increase in clinical POCUS scans (from 72 to 267 scans in the six months pre/post-intervention, p<0.001). We demonstrated that physicians, once trained, can use POCUS more often for medical diagnostic purposes clinically at the patient’s bedside.^32–34^ In contrast, this study evaluated an ultrasound-guided *procedural competency* simulation model as a standardized, reproducible model for future dissemination. The clinical NBs remained low after the intervention (21 pre- to 15 post-intervention), therefore additional instructional or reinforcement steps may be needed to bolster procedural education and translation into clinical practice.

### Limitations

Study limitations include the single center study design with a small sample size as a pilot study, which limits generalizability. Ultrasound training sessions were performed by the ultrasound fellow with the study PI present, and sometimes additional ultrasound faculty were teaching, which can create variability but mirrors realistic clinical teaching. All post-technical assessments were performed by the ultrasound fellow to limit variability, reinforce reliability, and limit bias by not involving the study PI. The lack of pre-intervention knowledge and technical skill assessments limits change comparison from pre- to post-intervention, but we included a 3-month post-intervention follow up measurement that suggested knowledge and skill retention. The post-intervention time point at three months could be repeated at six months or greater to determine prolonged sustainability. Also, qualitative analysis via semi-structured interviews or focus groups could further improve the intervention implementation and uptake by participants. The clinical POCUS image quality assurance scores are less useful in procedural POCUS because they are graded on a yes/no scale rather than the 5-point scale used for diagnostic exams. Thus, additional review by ultrasound experts with added clinical outcomes such as patient satisfaction with POCUS performance, pain scores, procedural complication rates, ED length-of-stay, and hospital admission vs discharge rates could be useful for detecting a difference in patient-centered outcomes.

### Future directions

Future multicenter studies with larger participant numbers and additional patient-centered clinical outcomes can detect if there is a true clinical impact in performing ultrasound-guided nerve blocks in EDs and other settings. Future studies can also expand this model to other ultrasound-guided nerve blocks or procedures and assess clinical outcomes such as patient pain, complications, and ED metrics.

## Conclusion

This low-fidelity POCUS simulation model increased knowledge, technical skills, and confidence scores in EM physicians and APPs performing simulated UG-SANB. Adaptation and testing of this model as a standardized, reproducible training method for other ultrasound applications and procedures could improve EM provider clinical skills but needs additional implementation research to improve ED clinical NB performance.

## Supporting information

S1 STROBE

S3 SANB knowledge exam

S4 UGNB survey

S5 SANB survey2

S2 Image of nerve block set up

## Data Availability

All data produced in the present study are available upon reasonable request to the authors.

## Notes

### Competing Interest Statement

The authors have declared no competing interest.

### Funding Statement

This grant was funded by an SAEMF Education Project Grant #RE2022-0000000349. The funders had no role in the design or execution of the study.

### Author Declarations

This study was determined exempt and consent was waived by the Duke University Health System Institutional Review Board (Pro00114989).

