## Supplementary material for "Emergency Physician Ultrasound-guided Nerve Block Training Simulation Assessment: a Prospective Cohort Study": S3 SANB knowledge exam

RT version of modified checklist for SANB:

Modified Cheung checklist for technical skills exam:

Passage of exam requires visualization of the needle tip at the fascial plane between the serratus anterior and latissimus dorsi muscles along with the presence of 10 or more of the following behaviors (adapted from Wong et al).

1. Optimizes patient positioning (supine for anterior approach or in lateral decubitus and turned away from the proceduralist for posterior approach)
2. Appropriately positions the ultrasound machine (US machine should be less than 45° off the line of sight from needle insertion)
3. chooses the correct transducer (linear)
4. chooses an appropriate depth, gain, and focal zone for needle visualization
5. uses Doppler to rule out vascular structures prior to needle advancement
6. uses stable grip to control the ultrasound probe (part of hand/finger should be anchored on the patient to stabilize the probe)
7. uses appropriate probe orientation (probe marker points to the user’s left and corresponds to the left side of the screen when the marker is on the left side of the screen)
8. positions the probe appropriately (transverse position on the lateral chest wall overlying the posterior axillary line at the level of the 5^th^ rib)
9. has an efficient and appropriate needle pathway (e.g for an anterior approach, the needle enters anterior to the probe and is directed posteriorly/inferiorly along the lateral chest wall)
10. maintains the image of the serratus anterior plane target throughout the procedure
11. maintains needle tip visualization during needle advancement, and immediately stops needle advancement if visualization is lost
12. targets spread of local anesthetic at the fascial plane to achieve hydro dissection between the latissimus dorsi and serratus anterior muscles
13. recognizes that the needle tip is adjacent to the nerve without touching the nerve itself
14. asks for syringe aspiration prior to injection to prevent inadvertent blood vessel injection

RT version of knowledge exam for SANB:

1. Which of the following is an indication for serratus anterior plane nerve block (SANB)? Check all that apply.
2. Which of the following is a contraindication for an SANB?
3. What nerves are reliably anesthetized by the SANB?
4. What is the most likely potential complication of the SANB, of the below options?
5. Approximately what are the maximum therapeutic doses of lidocaine without epinephrine and bupivacaine without epinephrine, respectively?
6. The most important concept for avoidance of complications from ultrasound guided nerve blocks is:
7. What organ system is the first to display signs of lidocaine and bupivacaine toxicity?
8. Lidocaine and bupivacaine toxicity (LAST) can cause severe refractory cardiac dysrhythmias. In addition to standard ACLS, what is the correct treatment for LAST?
9. Immediately after performing an SANB for your patient, she maintains intact sensation over the entire affected chest wall. Your best next step is:
10. What is the volume of bupivacaine anesthestic injection for a serratus anterior plane block?
11. The safest target for the SANB is located:

Answers:

1. Indication for SANB

a. shoulder dislocation

b. Posterior rib fractures

**c. prior to chest tube placement for pneumothorax**

**d. Anterior or lateral thoracic rib fractures**

2. Contraindication for SANB

a. coagulopathy

**b. soft tissue infection in the injection area**

c. recent thoracic surgery

d. patients with severe pulmonary disease

3. What nerves are innervated?

**a. lateral cutaneous branches of the thoracic intercostal nerves**

b. dorsal and ventral rami of the spinal nerves

c. thoracic nerve root

d. sympathetic ganglion

4. Most likely complication

a. local anesthetic systemic toxicity (LAST)

**b. pneumothorax**

c. acute nerve injury

d. pericardial puncture and effusion

5. What is the max dose of lidocaine/bupivacaine without epi?

a. maximum dose is 2 mg/kg of bupivacaine without epi, 2 mg/kg lidocaine without epi

b. maximum dose is 5 mg/kg of bupivacaine without epi, 5 mg/kg lidocaine without epi

**c. maximum dose is 2 mg/kg of bupivacaine without epi, 5 mg/kg lidocaine without epi**

d. maximum dose is 5 mg/kg of bupivacaine without epi, 2 mg/kg lidocaine without epi

6. What is the best way to prevent any complications when performing NB?

a. negative aspiration for blood prior to anesthetic injection

**b. always maintain the needle tip in view**

c. inject anesthetic as close to the nerve as possible to maximize analgesic effects

d. only inject 5ml of anesthetic so that you do not cause systemic toxicity

7. What system shows the first signs of LAST?

**a. nervous system**

b. cardiac system

c. pulmonary system

d. GI system

8. What is the appropriate treatment for LAST?

a. 0.5mg intramuscular epinephrine

**b. intravenous lipid emulsion 20% therapy, initial dose 1 to 2 g/kg/d**

c. 2g IV magnesium sulfate

d. 50mEq sodium bicarbonate

9. What do you do if the patient is still having pain after you performed your block?

a. inject more anesthetic into the same space

b. inject more anesthetic deep to the muscle

**c. wait 5-10 minutes**

d. give the patient IV morphine as the block failed

10. What is volume of bupivacaine without epi used for SANB?

a. 5 ml of 0.25% bupivacaine without epinephrine

**b. 30ml of 0.25% bupivacaine without epinephrine**

c. 5ml of 0.25% bupivacaine without epinephrine at the injection level plus 5ml at the rib above and below

d. 15 ml of 0.25% bupivacaine without epinephrine

11. What is the safest target for the SANB?

**a. at the fascial plane between the latissimus dorsi and serratus anterior muscles**

b. at the fascial plane between the pectoralis major and serratus anterior muscles

c. within the intercostal muscle between the lateral ribs

d. at the fascial plane between the trapezius and rhomboid muscles
