## Supplementary material for "Emergency Physician Ultrasound-guided Nerve Block Training Simulation Assessment: a Prospective Cohort Study": S4 UGNB survey

RT Pre/post-workshop survey

Please complete the survey. Your responses will be kept confidential.

Name: _________________

NetID: _____

Age: _____

Gender: (Male, Female, Nonbinary, or prefer not to answer)

Ethnicity: (Hispanic/Latino/x/Spanish origin or Non-Hispanic/Latino/x/Spanish origin, or prefer not to answer)

Race: (American Indian or Alaska Native, Asian, Black or African American, Native Hawaiian or Other Pacific Islander, White, other, or prefer not to answer)

-What is your level of training? (PGY1, 2, 3, fellow, attending physician, APP)

-Have you completed a fellowship? If so, which? Yes	No	_____________________

-Number of years in clinical practice working in the emergency department (including residency/fellowship)? _________

-If you are a resident, how long ago was your ultrasound rotation?

0-3 months, 3-6 months, 6-12 months, >12 months, N/A

--What forms of ultrasound training have you had to date? (Please select all that apply)

[ ] None

[ ] Podcasts, online blogs, FOAMed

[ ] Online ultrasound course

[ ] In-person ultrasound course or conference (approximately 1-3 days)

[ ] Designated ultrasound rotation or elective (approximately 1 month)

[ ] Bedside teaching and didactics throughout medical school

[ ] Bedside teaching and didactics throughout residency

[ ] Ultrasound fellowship training

[ ] Other (please specify) _____________________________________________________

--How many point-of-care ultrasound exams have you personally performed in medical school and residency?

a) 0-50

b) 51-100

c) 101-150

d) >150

--How confident do you feel performing ANY clinical nerve block with ultrasound-guidance **before/after** the intervention?

Very unconfident, unconfident, neutral, confident, very confident

--How confident do you feel performing serratus anterior clinical nerve block with ultrasound-guidance **before/after** the intervention?

Very unconfident, unconfident, neutral, confident, very confident

--How confident do you feel teaching others how to perform ANY clinical nerve block with ultrasound-guidance **before/after** the intervention?

Very unconfident, unconfident, neutral, confident, very confident

--How confident do you feel teaching others how to perform serratus anterior clinical nerve block with ultrasound-guidance **before/after** the intervention?

Very unconfident, unconfident, neutral, confident, very confident

--Do you believe that POCUS is a useful tool for medical decision making in the ED?

Strongly disagree, disagree, neutral, agree, strongly agree

--Do you believe it is important for residents to be skilled in using POCUS?

Strongly disagree, disagree, neutral, agree, strongly agree

Post-Course Evaluation

How often have you performed ANY clinical nerve block prior to the workshop?

a) Often (>1 time per week)

b) Somewhat often (1-2 times per month)

c) Occasional (<1 time per month)

d) Rarely (1-2 months per year)

e) Never

How often have you performed clinical serratus anterior plane nerve blocks (SANB) prior to the workshop?

a) Often (>1 time per week)

b) Somewhat often (1-2 times per month)

c) Occasional (<1 time per month)

d) Rarely (1-2 months per year)

e) Never

How often do you think you will perform ANY clinical nerve block after completing the workshop?

a) Often (>1 time per week)

b) Somewhat often (1-2 times per month)

c) Occasional (<1 time per month)

d) Rarely (1-2 months per year)

e) Never

How often do you think you will perform clinical serratus anterior plane nerve blocks (SANB) after completing the workshop?

a) Often (>1 time per week)

b) Somewhat often (1-2 times per month)

c) Occasional (<1 time per month)

d) Rarely (1-2 months per year)

e) Never

Please answer the following questions about the nerve block workshop.

(Strongly disagree, disagree, neutral, agree, strongly agree)

Implementation evaluation of UG-SANB education and simulation workshop for EM physicians

Acceptability of Intervention Measure (AIM):

1.	I approve of the education and simulation workshop.

2.	The simulation workshop teaching format is appealing to me.

3.	I like the education and simulation workshop timeframe.

4.	I like the hands-on aspect of the simulation model.

Intervention Appropriateness Measure (IAM):

1.	The simulation workshop structure and format are fitting.

2.	The simulation workshop seems suitable to my training level.

3.	The simulation workshop seems applicable for use in clinical care.

4.	The education and simulation workshop are a good match as a simple and adaptable model for ultrasound procedural training.

Feasibility of Intervention Measure (FIM):

1.	The simulation workshop and assessments seem implementable and reproducible for other procedures.

2.	The simulation workshop is possible to complete in the given time.

3.	It is doable to attend and participate in the simulation workshop.

4.	The simulation model seems easy to use.

Please answer the following questions about the nerve block workshop.

(Strongly disagree, disagree, neutral, agree, strongly agree)

-The pre-workshop self-learning training materials assisted my learning.

-The workshop teaching format was effective.

-The content covered was appropriate to my skill level.

-I gained new knowledge and skills by participating in the workshop.

-I will apply the knowledge and skills from the workshop for clinical patient care.

-I will apply the knowledge and skills from the workshop to teach other learners.

-Overall, I am satisfied with the course.

-I would recommend this course to a colleague.

Which component of the nerve block workshop was useful to you? _______________

What could be improved for future training interventions? ______________
