## Supplementary material for "Emergency Physician Ultrasound-guided Nerve Block Training Simulation Assessment: a Prospective Cohort Study": S5 SANB survey2

Supplemental File 5: Participant demographics from pre-surveys

|  | Pre-Intervention |
| --- | --- |
|  | (N = 63) |

| **Age (years)** |  |
| --- | --- |
| Mean (SD) | 33.0 (7.7) |
| Median (Q1, Q3) | 30.0 (28.0, 34.0) |
| Min, Max | 26.0, 58.0 |
| **Age (years)** |  |
| <30 | 26 (41.3%) |
| 30-40 | 23 (36.5%) |
| 41-50 | 6 (9.5%) |
| 51+ | 3 (4.8%) |
| Missing | 6 |
| **Gender** |  |
| Male | 38 (60.3%) |
| Female | 24 (38.0%) |
| Missing | 1 |
| **Ethnicity** |  |
| Hispanic/Latino/Spanish origin | 47 (74.6%) |
| Non-Hispanic/Latino/Spanish origin | 9 (14.2%) |
| Missing | 6 |
| **Race** |  |
| Asian | 10 (15.9%) |
| Black or African American | 7 (11.1%) |
| White | 37 (58.7%) |
| Other | 1 (1.6%) |
| Missing | 8 |
| **What is your level of training?** |  |
| Attending physician | 19 (30.2%) |
| Resident* | 31 (49.2%) |
| PGY3+ | 8 (22.2%) |
| PGY2 | 12 (33.3%) |
| PGY1 | 11 (30.6%) |
| APP | 13 (20.6%) |
| Missing | 0 |
| **Completed a fellowship**** | 10 (15.9%) |
| Missing | 0 |
| **Number of years in clinical practice working in the emergency department (including residency and fellowship)** |  |
| Mean (SD) | 5.5 (7.1) |
| Median (Q1, Q3) | 2.5 (1.0, 6.5) |
| Min, Max | 0.33, 30.0 |
| **Number of POCUS personally performed in residency training** |  |
| >150 | 26 (41.3%) |
| 101-150 | 12 (19.0%) |
| 51-100 | 7 (11.1%) |
| 0-50 | 17 (27.0%) |

*Percentages associated with PGY1, PGY2, and PGY3+ are calculated with a denominator of all residents

**Fellowships completed: health services research, emergency medical services, sports medicine, hyperbaric and undersea medicine, geriatric emergency medicine, emergency ultrasound

**Legend**: SD (standard deviation), PGY (post-graduate year), APP (Advanced Practice Provider), POCUS (point-of-care-ultrasound)
