## Supplementary figures and images for "Emergency Physician Ultrasound-guided Nerve Block Training Simulation Assessment: a Prospective Cohort Study"

### S2 Image of nerve block set up

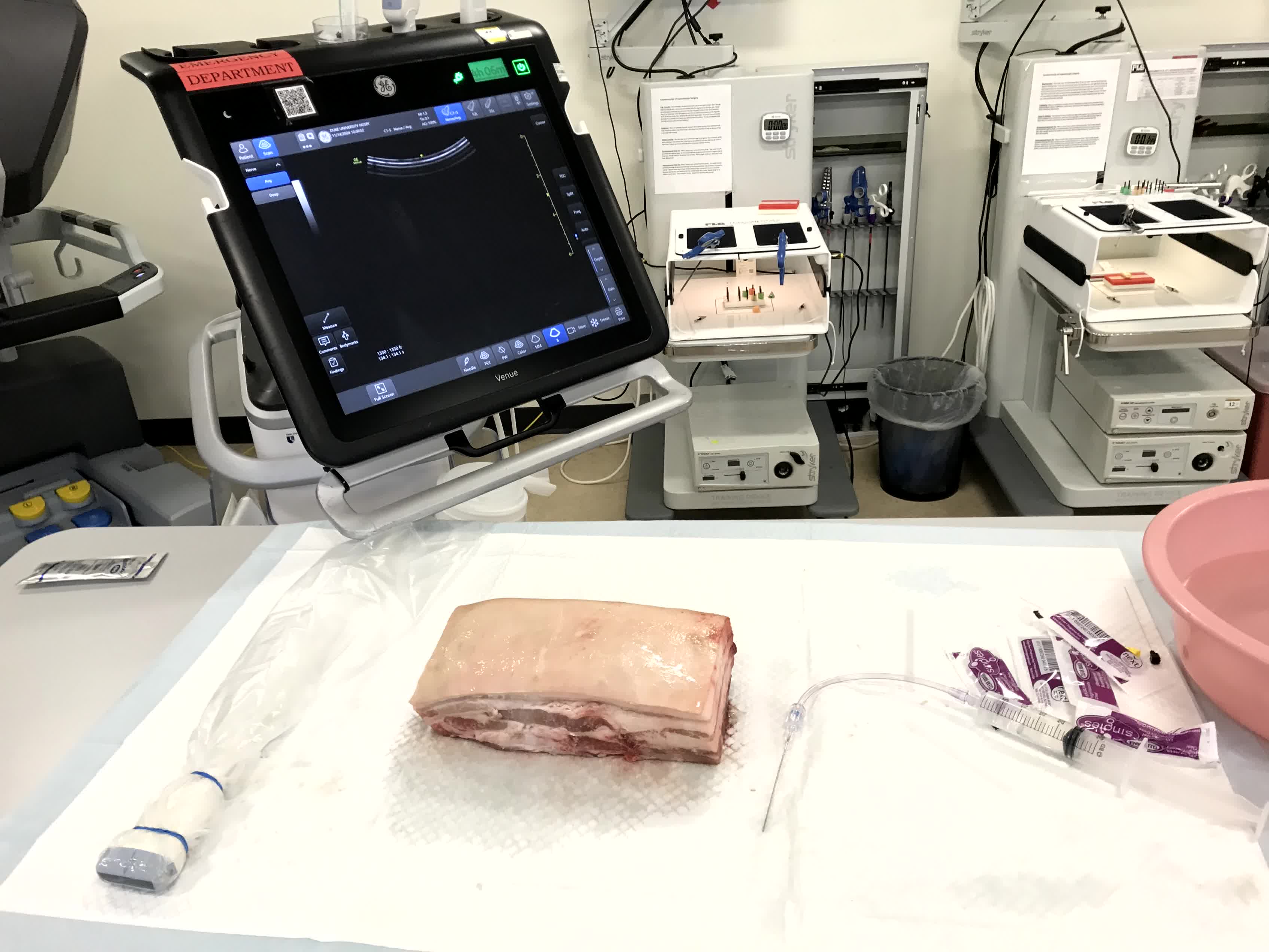
